# Development of an electronic frailty index for predicting mortality in patients undergoing transcatheter aortic valve replacement using machine learning

**DOI:** 10.1101/2020.12.23.20248770

**Authors:** Yiyi Chen, Jiandong Zhou, Sharen Lee, Tong Liu, Sandeep S Hothi, Ian Chi Kei Wong, Qingpeng Zhang, Gary Tse, Yan Wang

## Abstract

**Background:** Electronic frailty indices can be useful surrogate measures of frailty. We assessed the role of machine learning to develop an electronic frailty index, incorporating demographics, baseline comorbidities, healthcare utilization characteristics, electrocardiographic measurements, and laboratory examinations, and used this to predict all-cause mortality in patients undergoing transaortic valvular replacement (TAVR).

**Methods:** This was a multi-centre retrospective observational study of patients undergoing for TAVR. Significant univariate and multivariate predictors of all-cause mortality were identified using Cox regression. Importance ranking of variables was obtained with a gradient boosting survival tree (GBST) model, a supervised sequential ensemble learning algorithm, and used to build the frailty models. Comparisons were made between multivariate Cox, GBST and random survival forest models.

**Results:** A total of 450 patients (49% females; median age at procedure 82.3 (interquartile range, IQR 79.0-86.0)) were included, of which 22 died during follow-up. A machine learning survival analysis model found that the most important predictors of mortality were APTT, followed by INR, severity of tricuspid regurgitation, cumulative hospital stays, cumulative number of readmissions, creatinine, urate, ALP, and QTc/QT intervals. GBST significantly outperformed random survival forests and multivariate Cox regression (precision: 0.91, recall: 0.89, AUC: 0.93, C-index: 0.96, and KS-index: 0.50) for mortality prediction.

**Conclusions:** An electronic frailty index incorporating multi-domain data can efficiently predict all-cause mortality in patients undergoing TAVR. A machine learning survival learning model significantly improves the risk prediction performance of the frailty models.

## Introduction

Aortic stenosis (AS), reduction in the effective orifice area of the semilunar cardiac valve at the interface of the left ventricle and the systemic arterial circulation, is a significant medical problem globally ^1^. AS has a prevalence of 2-9% of the population over 65 years and 4% in those who are over 85 ^2^. It is associated with aging and confers a poor prognosis ^3^. There are different types of operations for the treatment of AS. Of these, transcatheter aortic valve replacement (TAVR) is a less invasive alternative to surgical aortic valve replacement for patients with severe, symptomatic AS, especially for those who are at high risk or intermediate risk of adverse events, e.g. those with a high number of comorbidities ^4^. Moreover, recent work has found that TAVR in low-risk patients is noninferior to surgical management ^5^.

TAVR has been shown to have a satisfactory efficacy and safety and recommendations by guidelines from various international societies ^6, 7^. However, recent studies have found that frailty is a common finding in AS patients and is associated with increased mortality. Approximately half of patients who undergo TAVR die within four years of the procedure ^8^. It is therefore crucial to weigh up risks and benefits before when offering TAVR to patients, in particular, trying to select those in whom the procedure is likely to confer greater gains in symptoms and prognosis, and identifying those who may not benefit, or indeed suffer harm, following TAVR.

Machine learning techniques have been widely applied in medical research. Specifically, a gradient boosting survival tree model has recently been explored as an efficient method for diagnosing coronary artery disease ^9^. In this territory-wide study, we tested the hypothesis that an electronic frailty index incorporating demographics, baseline comorbidities, healthcare utilization characteristics, electrocardiographic measurements, and laboratory examinations using a gradient boosting approach can improve risk prediction for all-cause mortality.

## Methods

### Study design

This study was approved by The Joint Chinese University of Hong Kong - New Territories East Cluster Clinical Research Ethics Committee. This retrospective study included patients undergoing TAVR. Patients were identified from the Clinical Data Analysis and Reporting System (CDARS), a healthcare database that integrates patient information across all 43 publicly funded hospitals and their associated ambulatory and primary care facilities in Hong Kong to establish comprehensive medical records. The available information includes demographics, clinical characteristics, disease diagnoses, laboratory examinations, drug prescription details, and admission statistics.

### Data extraction and variables

The following data were extracted: 1) Baseline characteristics of gender, age at TAVR, age at first presentation with AS, TR severity, AR severity, MR severity, PR severity, complete recovery status, INR on the day of TAVR procedure; 2) baseline comorbidities including bradyarrhythmia, atrial fibrillation/flutter, tachyarrhythmia, diabetes mellitus, hypertension, hyperlipidaemia, respiratory diseases, kidney diseases, endocrine disorders (other than diabetes mellitus) and gastrointestinal diseases using corresponding ICD-9 codes; 3) ECG measurements of ventricular rate, P-wave duration (PWD), PR interval, QRS duration, QT interval, corrected QT interval (QTc), P-wave axis, QRS axis, T-wave axis, R-wave amplitude in V5, and S-wave amplitude in V1; 4) healthcare utilization characteristics before TAVR presentation: cumulative hospital stay, cumulative number of hospital readmissions, and number of emergency readmissions (within 28 days of discharge); 5) laboratory tests: complete blood count, liver function tests, renal function tests. Details of ICD codes used for comorbidity identification are provided in the **Supplementary Appendix**.

### Primary outcome and statistical analysis

The primary outcome was all-cause mortality. Descriptive statistics were presented for the overall cohort and categorized based on mortality status. Continuous variables were presented as median (95% confidence interval [CI] or interquartile range [IQR]) and categorical variables were presented as counts (%). The Mann-Whitney U test was used to compare continuous variables. The χ^2^ test with Yates’ correction was used for 2×2 contingency data, and Pearson’s χ^2^ test was used for contingency data for variables with more than two categories. To evaluate the significant prognostic risk factors associated with disease group status and primary outcomes, univariate Cox regression models were used with adjustments based on baseline characteristics. Significant univariable predictors were used as inputs in a multivariate Cox regression model to avoid overfitting. Hazard ratios (HRs) with corresponding 95% CIs and P values were reported accordingly. All significance tests were two-tailed and considered statistically significant if P values were 0.05. Data analyses were performed using RStudio software (Version: 1.1.456) and Python (Version: 3.6). Simulations were performed were using a 15-inch MacBook Pro with 2.2 GHz Intel Core i7 Processor and 16 GB RAM (Hong Kong, China).

### Development of a gradient boosting survival tree model

Survival analysis is a statistical method to deal with lifetime data, where the outcome is the time to occurrence of an event of interest, such as mortality. The most widely applied survival analysis model in biostatistics is the Cox proportional hazards model ^10^. Gradient boosting, a class of machine learning methods, was developed based on the concept that a tree-based model after being sequentially combined with previous weak models (e.g. decision trees) in a stage-wise way can generate superior predictions for survival and other outcomes. Tree-structured survival models such as survival trees ^11^ and random survival forests ^12^ have been used to determine survival probabilities in medical studies.

This inspired us to use a nonparametric ensemble tree model called a gradient boosting survival tree (GBST) that extends the survival tree models with the concept of gradient boosting ^13^. GBST optimizes the survival probability of each time period simultaneously and therefore is able to significantly reduce the overall prediction error of a survival tree. In this study, GBST was used for mortality risk prediction of patients undergoing TAVR. A tree-structure based approach for ranking the importance value of different variables was used, to construct a machine learning based, electronic frailty index for predicting mortality outcome. To examine the GBST performance of survival risk discrimination and compare it with baseline models of random survival forests and a multivariate Cox regression model, we adopted a five-fold cross validation approach. The concordance index (C Index) proposed by Harrell et al. (1982) was used to measure the goodness of fit for the survival model, as the statistic provides a global assessment of the model for the continuous event. The territory-wide cohort dataset also guarantees the c-Index not to be limited by censoring. We applied the c-index introduced above as well as a precision, recall, Kolmogorov-Smirnov (KS) index and the area under the receiver operating characteristics curve (AUC) to measure the goodness of model fit. The R packages, gbm (Version 2.1.5), randomForestSRC (Version 2.9.3), survival (Version 2.42-3) and ggplot2 (Version 3.3.2), were used to generate the mortality prediction results.

## Results

### Baseline cohort characteristics

The baseline characteristics of this cohort of patients undergoing TAVR are shown in **Table 1**. A total of 450 patients were included (female 49%; mean age at time of procedure: 82.3 (IQR 79.0-86.0) years; median age at first hospital presentation:79 (IQR: 74.0-83.0)) years. A total of 22 deaths (32% female) occurred during follow-up. The Kaplan-Meier survival curve of patients undergoing TAVR is shown in **Figure 1**.

**Table 1.** Descriptive statistics of the TAVR cohorts. ***:<0.001; **:<0.01;*:<0.05;.:< 0.1

| **Characteristics** | **All patients (n=450)**<br>**Median (IQR) or count (%)** | **Death (n=22)**<br>**Median (IQR) or count (%)** | **Alive (n=428)**<br>**Median (IQR) or count (%)** | **P** |
| --- | --- | --- | --- | --- |
| **Demographics** |  |  |  |  |
| Female sex | 221(49.11%) | 7(31.81%) | 214(50.00%) | 0.0026** |
| Age at TAVR | 82.3(79.0-86.0) | 83.8(81.0-87.0) | 82.1(79.0-86.0) | 0.261 |
| Age at first visit | 79(74.0-83.0) | 81(78.0-84.0) | 79(74.0-83.0) | 0.017* |
| TR Severity |  |  |  |  |
| None/trivial | 174(38.66%) | 7(31.81%) | 167(39.01%) | 0.0016** |
| Mild | 177(39.33%) | 4(18.18%) | 173(40.42%) | <0.0001*** |
| Moderate | 78(17.33%) | 6(27.27%) | 72(16.82%) | 0.66 |
| Severe | 21(4.66%) | 5(22.72%) | 16(3.73%) | 0.021* |
| AR Severity |  |  |  |  |
| None/trivial | 167(37.11%) | 9(40.90%) | 158(36.91%) | 0.002** |
| Mild | 217(48.22%) | 9(40.90%) | 208(48.59%) | <0.0001*** |
| Moderate | 60(13.33%) | 3(13.63%) | 57(13.31%) | <0.0001*** |
| Severe | 6(1.33%) | 1(4.54%) | 5(1.16%) | 0.121 |
| MR Severity |  |  |  |  |
| None/trivial | 140(31.11%) | 4(18.18%) | 136(31.77%) | 0.172 |
| Mild | 222(49.33%) | 14(63.63%) | 208(48.59%) | <0.0001*** |
| Moderate | 75(16.66%) | 2(9.09%) | 73(17.05%) | <0.0001*** |
| Severe | 13(2.88%) | 2(9.09%) | 11(2.57%) | 0.012* |
| PR Severity |  |  |  |  |
| None/trivial | 167(37.11%) | 9(40.90%) | 158(36.91%) | 0.021* |
| Mild | 217(48.22%) | 9(40.90%) | 208(48.59%) | <0.0001*** |
| Moderate | 60(13.33%) | 3(13.63%) | 57(13.31%) | <0.0001*** |
| Severe | 6(1.33%) | 1(4.54%) | 5(1.16%) | 0.123 |
| Complete recovery | 129(28.66%) | 0(0.00%) | 129(30.14%) | <0.0001*** |
| INR | 1.07(1.0-1.19);n=450 | 1.32(1.07-2.385);n=22 | 1.07(1.0-1.17);n=428 | <0.0001*** |
| **Comorbidities** |  |  |  |  |
| Bradyarrhythmia | 23(5.11%) | 1(4.54%) | 22(5.14%) | <0.0001*** |
| Tachyarrhythmia | 34(7.55%) | 0(0.00%) | 34(7.94%) | <0.0001*** |
| Diabetes/Hypertension | 318(70.66%) | 17(77.27%) | 301(70.32%) | <0.0001*** |
| Hyperlipidaemia | 143(31.77%) | 5(22.72%) | 138(32.24%) | <0.0001*** |
| Atrial fibrillation/flutter | 156(34.66%) | 11(50.00%) | 145(33.87%) | 0.0016** |
| Respiratory | 269(59.77%) | 16(72.72%) | 253(59.11%) | <0.0001*** |
| Kidney | 174(38.66%) | 12(54.54%) | 162(37.85%) | <0.0001*** |
| Endocrine | 23(5.11%) | 1(4.54%) | 22(5.14%) | 0.0172* |
| Gastrointestinal | 249(55.33%) | 9(40.90%) | 240(56.07%) | 0.016* |
| **ECG Measurements** |  |  |  |  |
| Ventricular Rate | 79(68.0-93.0);n=375 | 95(78.0-109.0);n=17 | 78(68.0-92.0);n=358 | 0.172 |
| PWD | 119(109.0-127.0);n=137 | 89.5(90.0-90.0);n=2 | 120(109.0-127.0);n=135 | <0.0001*** |
| PR interval | 173(154.0-196.0);n=290 | 175.5(146.0-190.0);n=12 | 173(154.0-196.0);n=278 | 0.187 |
| QRS | 98(90.0-110.0);n=375 | 95(90.0-106.0);n=17 | 98(90.0-110.0);n=358 | 0.23 |
| QT | 390(358.0-424.0);n=375 | 379(341.0-402.0);n=17 | 391(360.0-424.0);n=358 | 0.012* |
| QTc | 439(420.0-459.0);n=376 | 449(433.0-471.0);n=17 | 438(419.0-459.0);n=359 | 0.611 |
| P-wave axis | 54(30.0-72.0);n=329 | 49(32.0-94.0);n=11 | 54(30.0-72.0);n=318 | 0.341 |
| QRS axis | 45(22.0-70.0);n=375 | 54(20.0-70.0);n=17 | 45(22.0-70.0);n=358 | 0.62 |
| T-wave axis | 71(39.0-115.0);n=372 | 61(30.0-131.0);n=15 | 72(39.0-115.0);n=357 | 0.0023** |
| R-wave amp in V5 | 1.87(1.27-2.44);n=236 | 1.5(1.0-2.0);n=4 | 1.87(1.28-2.44);n=232 | 0.102 |
| S-wave amp in V1 | 1(1.0-2.0);n=235 | 0.8(0.0-1.0);n=4 | 0.95(0.526-1.535);n=231 | 0.081 |
| **Hospitalization characteristics** |  |  |  |  |
| Cumulative LOS | 18(6.0-48.0);n=395 | 53(10.0-121.0);n=19 | 18(6.0-46.0);n=376 | <0.0001*** |
| Cumulative no. of admissions | 9(4.0-15.0);n=395 | 12(4.0-23.0);n=19 | 9(4.0-15.0);n=376 | 0.0267* |
| Emergency no. of readmission | 2(1.0-5.0);n=155 | 2.5(1.0-6.0);n=12 | 2(1.0-4.0);n=143 | 0.261 |
| **CBC examinations** |  |  |  |  |
| MCV, fL | 90.9(87.0-94.0);n=444 | 91.4(88.0-96.0);n=22 | 90.9(87.0-94.0);n=422 | 0.7512 |
| Basophil, x10<sup>9</sup>/L | 0.02(0.01-0.04);n=393 | 0.02(0.01-0.025);n=17 | 0.02(0.01-0.04);n=376 | 0.97 |
| Eosinophil, x10<sup>9</sup>/L | 0.16(0.1-0.27);n=442 | 0.125(0.0745-0.275);n=22 | 0.17(0.1-0.27);n=420 | 0.781 |
| HbA1C, g/dL | 12.2(11.0-14.0);n=444 | 12(11.0-13.0);n=22 | 12.2(11.0-14.0);n=422 | 0.0561 |
| Lymphocyte, x10<sup>9</sup>/L | 1.5(1.1-1.975);n=442 | 1.08(0.81-1.48);n=22 | 1.5(1.0-2.0);n=420 | <0.0001*** |
| Blast, x10<sup>9</sup>/L | 0.056(0.0-0.1);n=139 | 0.045(0.0-0.065);n=12 | 0.054(0.0-0.1);n=127 | 0.0281* |
| Metamyelocyte, x10<sup>9</sup>/L | 0.11(0.075-0.19);n=16 | 0.09(0.055-0.165);n=4 | 0.115(0.08-0.225);n=12 | <0.0001*** |
| Monocyte, x10<sup>9</sup>/L | 0.45(0.35-0.63);n=442 | 0.5(0.34-0.6);n=22 | 0.4(0.0-1.0);n=420 | 0.2711 |
| APTT, sec | 12.1(11.0-14.0);n=439 | 15.2(12.0-19.0);n=22 | 12.1(11.0-14.0);n=417 | <0.0001*** |
| Neutrophil, x10<sup>9</sup>/L | 4.1(3.14-5.385);n=442 | 4.2(3.0-6.0);n=22 | 4.1(3.0-5.0);n=420 | 0.271 |
| WBC, x10<sup>9</sup>/L | 6.5(5.0-8.0);n=444 | 6.1(5.0-8.0);n=22 | 6.5(5.0-8.0);n=422 | 0.251 |
| MCH, g/dL | 33.5(33.0-34.0);n=444 | 33.9(33.0-34.0);n=22 | 33.5(33.0-34.0);n=422 | 0.325 |
| Myelocyte, x10<sup>9</sup>/L | 0.18(0.105-0.27);n=18 | 0.17(0.13-0.555);n=3 | 0.19(0.095-0.27);n=15 | 0.0182* |
| Platelet, x10<sup>9</sup>/L | 189.5(153.0-232.0);n=444 | 152.5(130.0-195.0);n=22 | 190(155.0-233.0);n=422 | <0.0001*** |
| Reticulocyte, x10<sup>9</sup>/L | 55.2(40.0-79.0);n=65 | 50.6(44.0-77.0);n=5 | 55.2(39.0-80.0);n=60 | <0.0001*** |
| RBC, x10<sup>12</sup>/L | 4.13(3.685-4.52);n=444 | 3.945(3.585-4.45);n=22 | 4.1(4.0-5.0);n=422 | <0.0001*** |
| HCT, L/L | 0.352(0.32-0.39);n=201 | 0.399(0.3805-0.4095);n=6 | 0.35(0.32-0.39);n=195 | 0.0207* |
| **LRFT examinations** |  |  |  |  |
| Potassium, mmol/L | 4.2(4.0-4.0);n=421 | 4.4(4.0-4.8);n=20 | 4.2(3.8-4.5);n=340 | <0.0001*** |
| Urate, mmol/L | 0.39(0.32-0.47);n=240 | 0.43(0.36-0.48);n=16 | 0.39(0.32-0.47);n=224 | 0.0016** |
| Albumin, g/L | 40(37.0-43.0);n=445 | 38.9(36.0-41.0);n=22 | 40(37.0-43.0);n=423 | 0.0121* |
| Urea, mmol/L | 6.4(5.0-8.0);n=445 | 6.6(6.0-10.0);n=22 | 6.4(5.0-8.0);n=423 | 0.0132* |
| Sodium, mmol/L | 140.1(138.0-142.0);n=360 | 140.7(139.0-143.0);n=20 | 140(138.0-142.0);n=340 | 0.271 |
| Protein, g/L | 72(68.0-76.0);n=400 | 73.2(67.0-76.0);n=20 | 72(68.0-76.0);n=380 | 0.835 |
| Creatinine, umol/L | 88.4(71.0-110.0);n=445 | 105.9(78.0-150.0);n=22 | 88(70.0-108.0);n=423 | <0.0001*** |
| ALP, U/L | 70(59.0-86.0);n=445 | 73.5(64.0-91.0);n=22 | 70(58.0-86.0);n=423 | 0.356 |
| Aspartate Transaminase, U/L | 26(21.0-34.0);n=260 | 24.8(22.0-32.0);n=8 | 26(21.0-34.0);n=252 | 0.161 |
| ALT, U/L | 18(13.0-24.0);n=442 | 14.5(12.0-19.0);n=22 | 18(13.0-24.0);n=420 | <0.0001*** |
| Bilirubin, umol/L | 10.4(8.0-14.0);n=445 | 13(9.0-19.0);n=22 | 10.3(8.0-14.0);n=423 | 0.0015** |

**Figure 1.**
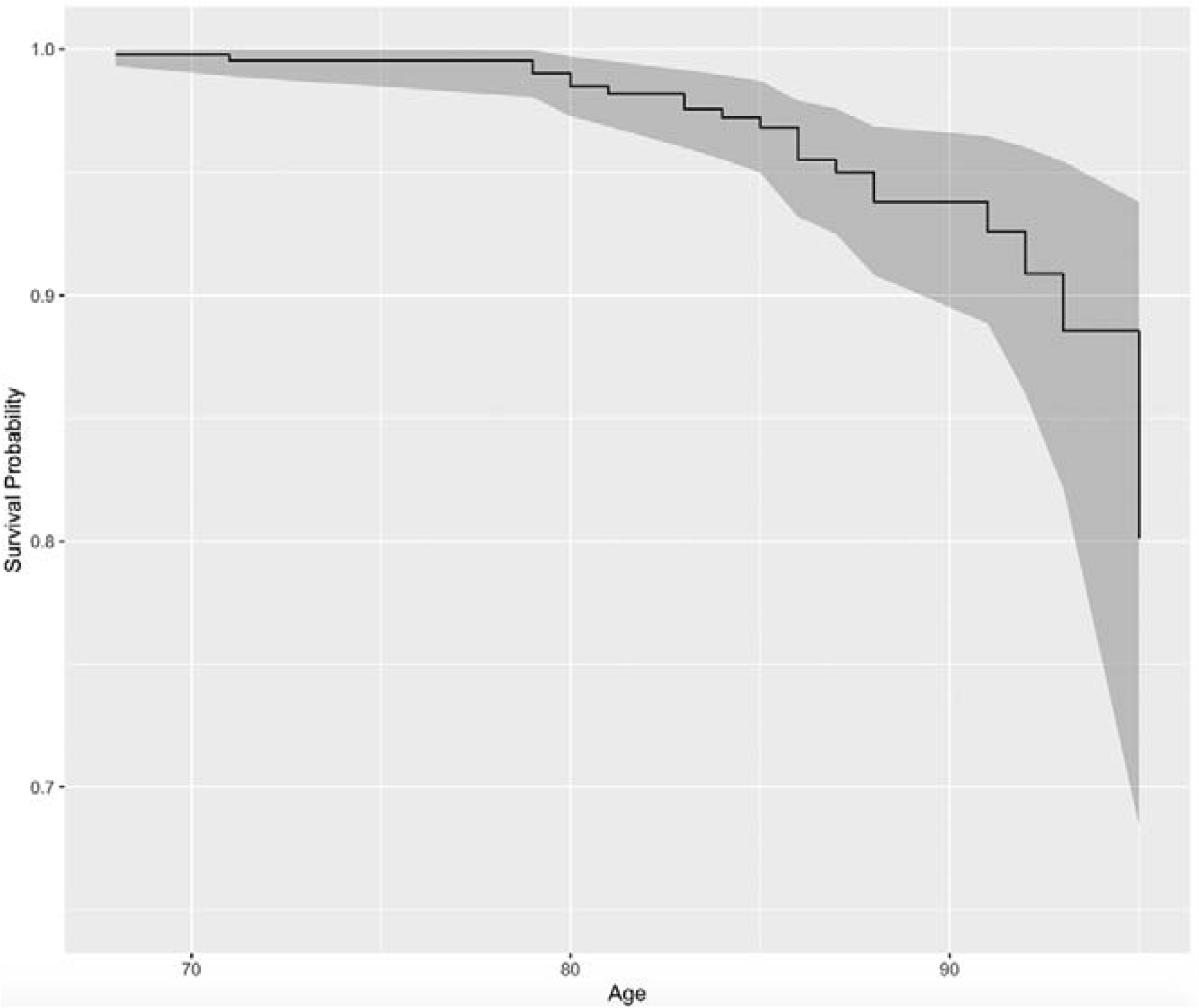
Kaplan-Meier survival curves of patients undergoing TAVR.

Further comparisons were made between the deceased and alive subgroups. Patients who died had an older age (83.8, IQR: [81.0-87.0] vs. 82.1, IQR: [79.0-86.0]) years, higher INR at procedure (1.32, IQR: 1.07-2.39 vs. 1.07, IQR: [1.0-1.17]). The presence of diabetes mellitus and hypertension were the most common comorbidities, followed by respiratory, gastrointestinal, and kidney diseases, atrial fibrillation/flutter, and hyperlipidaemia. In terms of healthcare utilization, those who died had a longer median in-hospital length-of-stay (53 days, IQR: [10.0-121.0] vs. 18 days, IQR: [6.0-46.0]), higher median number of hospital admissions (12, IQR: [4.0-23.0] vs. 9, IQR: [4.0-15.0]) and median number of emergency readmissions within 28 days after discharge (2.5, IQR: [1.0-6.0] vs. 2, IQR: [1.0-4.0]) before the TAVR procedure.

Regarding laboratory examinations, those who died had larger mean corpuscular volume (91.4 fL, IQR: [88.0-96.0]), higher mean corpuscular haemoglobin (33.9 x10^9/L, IQR: [33.0-34.0]), higher haematocrit (0.399, L/L, IQR: [0.3805-0.4095]), higher neutrophil count (4.2 x10^9/L, IQR: [3.0-6.0]), higher monocyte count (0.5 x10^9/L, IQR: [0.34-0.6]) and longer APTT (15.2 secs, IQR: [12.0-19.0]). By contrast, they had lower lower red blood cell (50.6 x10^12/L, IQR: [44.0-77.0]), white blood cell (median: 6.1 x10^9/L, IQR: [5.0-8.0]), eosinophil (0.125 x10^9/L, IQR: [0.0745-0.275]), lymphocyte (1.08 x10^9/L, IQR: [0.81-1.48]), blast (0.045 x10^9/L, IQR: [0.0-0.065]), metamyelocyte (0.09 x10^9/L, IQR: [0.055-0.165]) and myelocyte counts (0.17 x10^9/L, IQR: [0.13-0.555]) as well as lower platelet counts (152.5 x10^9/L, IQR: [130.0-195.0]) compared to those who remained alive (**Table 1**).

Moreover, they had higher potassium (4.4 mmol/L, IQR: [4.0-4.8]), urate (0.43 mmol/L, IQR: [0.36-0.48]), urea (6.6 mmol/L, IQR: [6.0-10.0]), protein (73.2 g/L, IQR: [67.0-76.0]), creatinine (105.9 umol/L, IQR: [78.0-150.0]), alkaline phosphatase (73.5 U/L, IQR: [64.0-91.0]) and bilirubin levels (13, umol/L, IQR: [9.0-19.0]), but lower sodium (140 mmol/L, IQR: [140.0-142.0]), albumin (38.9 g/L, IQR: [36.0-41.0]), aspartate transaminase (median: 24.8 U/L, IQR: [22.0-32.0]) as well as HbA1c levels (12 g/dL, IQR: [11.0-13.0]).

In terms of ECG measurements, patients who died had higher basal ventricular rate (95, IQR: [78-109] vs. 78 bpm, IQR: [68-92]), lower P-wave durations (90, IQR: [90-90] vs. 120 ms, IQR: [109-127]). PR interval, QTc interval and QRS axis were all significantly larger, whereas QRS duration, QT interval, P-wave axis, T-wave axis, R-wave amplitude in V5 and S-wave amplitude in V1 were lower in those who died compared to those who were alive (**Table 1**).

### Predictors of mortality and frailty model

Univariate Cox regression analysis was performed to identify significant predictors of all-cause mortality (**Table 2**). This identified severe tricuspid regurgitation (hazard ratio [HR]: 8.93, 95%CI: [3.22, 24.78], p<0.0001), international normalized ratio (HR: 2.74, 95% CI: [1.84, 4.09], p<0.0001), cumulative hospital length-of-stay (HR: 1.01, 95% CI: [1.00, 1.01], p=0.0008), aspartate transaminase (HR: 1.01, 95% CI: [0.98, 1.002], p=0.0002***), and bilirubin (HR: 1.02, 95% CI: [1.01, 1.02], p=0.0003***) as predictors of all-cause mortality. Subsequently, important variables (HR≥1) were included in a multivariate regression analysis (**Table 3**). Severe tricuspid regurgitation, INR, haematocrit and potassium were significant predictors after adjustment (P<0.001).

**Table 2.** Univariate regression analysis to identify the predictors of TAVR mortality. ***:<0.001; **:<0.01;*:<0.05;.:< 0.1

| **Characteristics** | **Hazard ratio (95% CI)** | **Z value** | **P** |
| --- | --- | --- | --- |
| **Demographics** |  |  |  |
| Female sex | 0.41[0.17, 1.01] | -1.947 | 0.0515 |
| Age at TAVR | 0.98[0.95, 1.02] | -1.099 | 0.272 |
| First presentation age | 0.99[0.95, 1.02] | -0.735 | 0.462 |
| TR Severity |  |  |  |
| None/trivial | 0.88[0.36, 2.16] | -0.278 | 0.781 |
| Mild | 0.38[0.13, 1.14] | -1.726 | 0.0843. |
| Moderate | 1.09[0.42, 2.86] | 0.175 | 0.861 |
| Severe | 8.93[3.22, 24.78] | 4.203 | 2.64E-05*** |
| AR Severity |  |  |  |
| None/trivial | 1.38[0.59, 3.23] | 0.732 | 0.464 |
| Mild | 0.64[0.27, 1.50] | -1.025 | 0.305 |
| Moderate | 1.10[0.33, 3.73] | 0.155 | 0.877 |
| Severe | 2.30[0.31, 17.27] | 0.81 | 0.418 |
| MR Severity |  |  |  |
| None/trivial | 0.62[0.21, 1.82] | -0.877 | 0.381 |
| Mild | 1.68[0.70, 4.00] | 1.164 | 0.245 |
| Moderate | 0.43[0.10, 1.83] | -1.144 | 0.253 |
| Severe | 2.79[0.64, 12.09] | 1.367 | 0.171 |
| PR Severity |  |  |  |
| None/trivial | 1.38[0.59, 3.23] | 0.732 | 0.464 |
| Mild | 0.64[0.27, 1.50] | -1.025 | 0.305 |
| Moderate | 1.10[0.33, 3.73] | 0.155 | 0.877 |
| Severe | 2.30[0.31, 17.27] | 0.81 | 0.418 |
| Complete recovery | - | - | - |
| **Comorbidities** |  |  |  |
| Bradyarrhythmia | 0.87[0.12, 6.47] | -0.136 | 0.892 |
| Tachyarrhythmia | - | - | - |
| Diabetes/Hypertension | 1.26[0.46, 3.44] | 0.457 | 0.647 |
| Hyperlipidaemia | 0.64[0.23, 1.73] | -0.884 | 0.377 |
| Atrial fibrillation/flutter | 7.23[0.96, 54.69] | 1.917 | 0.055 |
| Respiratory | 1.69[0.66, 4.33] | 1.095 | 0.274 |
| Kidney | 1.97[0.85, 4.57] | 1.579 | 0.114 |
| Endocrine | 0.85[0.11, 6.32] | -0.159 | 0.874 |
| Gastrointestinal | 0.49[0.21, 1.16] | -1.624 | 0.104 |
| **ECG Measurements** |  |  |  |
| Ventricular Rate | 1.02[1.00, 1.03] | 2.379 | 0.0174* |
| PWD | 0.94[0.88, 1.00] | -2.077 | 0.0378* |
| PR interval | 0.99[0.97, 1.01] | -1.059 | 0.29 |
| QRS | 1.00[0.98, 1.02] | 0.067 | 0.946 |
| QT | 1.00[0.99, 1.01] | -0.755 | 0.45 |
| QTc | 1.01[1.00, 1.02] | 2.434 | 0.0149* |
| P-wave axis | 1.00[1.00, 1.00] | -0.095 | 0.924 |
| QRS axis | 1.00[0.99, 1.02] | 0.475 | 0.635 |
| T-wave axis | 1.00[0.99, 1.01] | -0.013 | 0.99 |
| R-wave amplitude in V5 | 0.64[0.22, 1.87] | -0.823 | 0.411 |
| S-wave amplitude in V1 | 0.54[0.11, 2.55] | -0.778 | 0.436 |
| **Hospitalization characteristics** |  |  |  |
| Cumulative length-of-stay | 1.01[1.00, 1.01] | 2.618 | 0.001*** |
| Cumulative no. of admissions | 1.02[1.00, 1.04] | 2.161 | 0.031* |
| No. of emergency readmissions | 1.01[0.94, 1.10] | 0.337 | 0.736 |
| **CBC and clotting examinations** |  |  |  |
| MCV, fL | 1.04[0.97, 1.11] | 1.076 | 0.282 |
| Basophil, x10<sup>9</sup>/L | 0.80[0.02, 162.40] | -1.524 | 0.127 |
| Eosinophil, x10<sup>9</sup>/L | 0.75[0.10, 5.82] | -0.271 | 0.786 |
| HbA1C, g/dL | 0.97[0.74, 1.28] | -0.207 | 0.836 |
| Lymphocyte, x10<sup>9</sup>/L | 0.33[0.14, 0.79] | -2.482 | 0.013* |
| Blast, x10<sup>9</sup>/L | 0.08[0.00, 19481.00] | -0.407 | 0.684 |
| Metamyelocyte, x10<sup>9</sup>/L | 0.12[0.00, 428.20] | -0.502 | 0.616 |
| Monocyte, x10<sup>9</sup>/L | 0.62[0.09, 4.56] | -0.465 | 0.642 |
| APTT, sec | 1.02[1.00, 1.04] | 1.708 | 0.088 |
| Neutrophil, x10<sup>9</sup>/L | 0.99[0.83, 1.19] | -0.091 | 0.928 |
| WBC, x10<sup>9</sup>/L | 0.90[0.71, 1.13] | -0.933 | 0.351 |
| MCH, g/dL | 1.25[0.84, 1.85] | 1.094 | 0.274 |
| Myelocyte, x10<sup>9</sup>/L | 1.61[0.14, 18.84] | 0.38 | 0.704 |
| Platelet, x10<sup>9</sup>/L | 0.99[0.98, 1.00] | -2.164 | 0.0305* |
| Reticulocyte, x10<sup>9</sup>/L | 0.99[0.96, 1.02] | -0.543 | 0.587 |
| RBC, x10<sup>12</sup>/L | 0.66[0.32, 1.37] | -1.113 | 0.266 |
| HCT, L/L | 22.00[0.03, 31.00] | 1.584 | 0.113 |
| INR | 2.74[1.84, 4.09] | 4.955 | 7.23E-07*** |
| **LRFT examinations** |  |  |  |
| Potassium, mmol/L | 2.54[0.56, 5.12] | 0.526 | 0.091 |
| Urate, mmol/L | 2.23[0.11, 43.72] | 0.528 | 0.597 |
| Sodium, g/L | 1.25[0.35, 2.11] | 1.23 | 0.081 |
| Albumin, g/L | 0.95[0.86, 1.04] | -1.146 | 0.252 |
| Urea, mmol/L | 1.08[1.02, 1.14] | 2.649 | 0.008** |
| Protein, g/L | 1.02[0.95, 1.09] | 0.466 | 0.642 |
| Creatinine, umol/L | 1.00[1.00, 1.00] | 2.469 | 0.014* |
| ALP, U/L | 1.01[1.00, 1.02] | 2.335 | 0.020* |
| Aspartate Transaminase, U/L | 1.01[0.98, 1.002] | 3.721 | 0.0002*** |
| ALT, U/L | 1.00[1.00, 1.01] | 3.282 | 0.001** |
| Bilirubin, umol/L | 1.02[1.01, 1.02] | 3.59 | 0.0003*** |

**Table 3.**
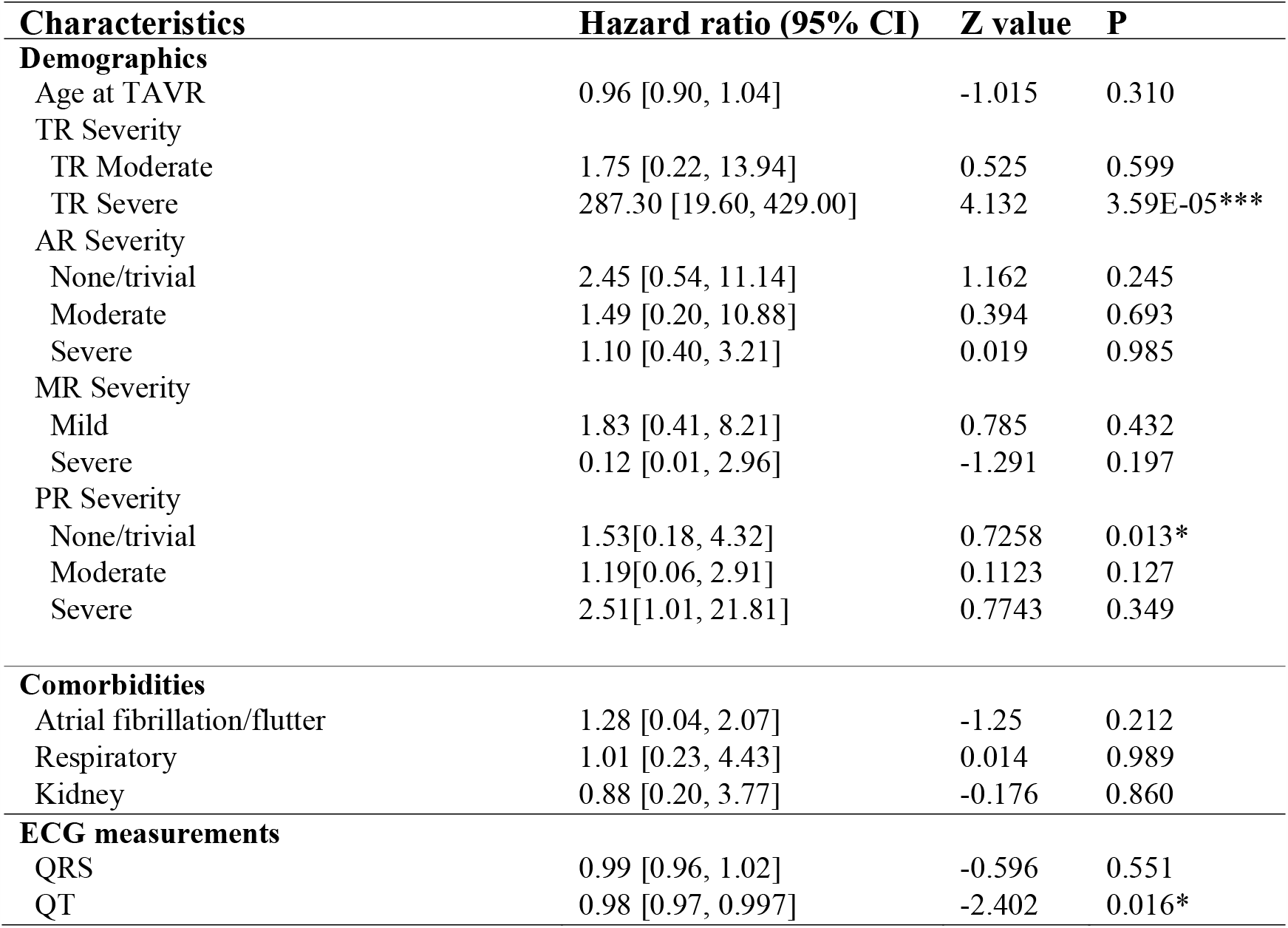
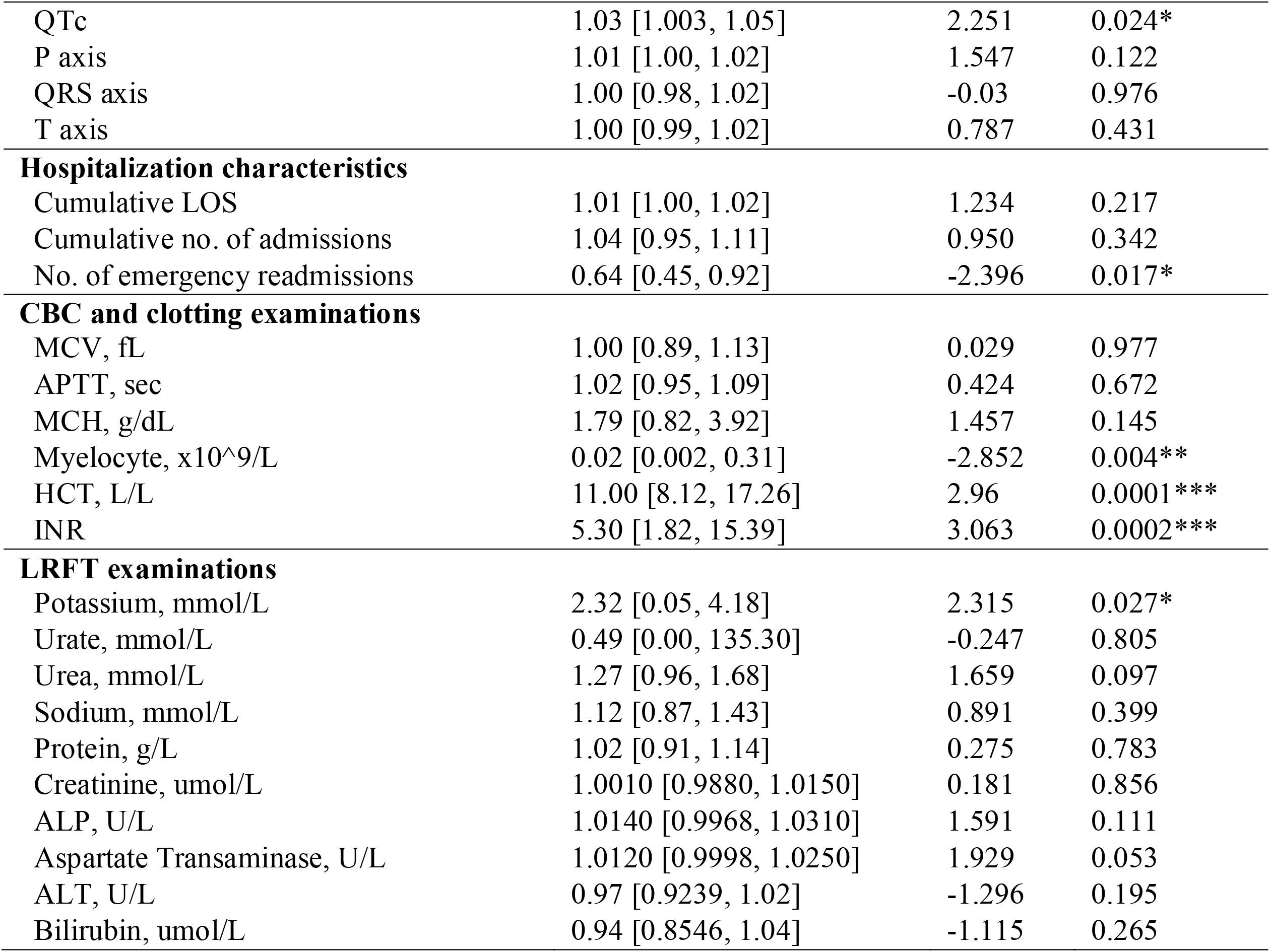
Multivariate regression analysis to identify the predictors of TAVR mortality. ***:<0.001; **:<0.01;*:<0.05;.:< 0.1

### Results of machine learning survival analysis and frailty score construction

The tree number in the GBST model was set to 320 according to the sensitivity analysis results (**Supplementary Figure 1**), where the association of Cox partial deviance versus the number of trees within the gradient boosting tree structure was depicted. The green line shows the validation deviance versus iteration number, with iteration almost stabilizing around the deviance value of 6.3 when one thousand trees were used. The black line shows the training error versus the iteration number. The optimal tree number was set according to the dashed blue line (validation).

With the input of important risk predictors identified by univariate Cox regression analysis, variable importance ranking is obtained by performing the introduced GBST model (**Supplementary Figure 2)** with the values shown in **Supplementary Table 2**. APTT showed the most important strength, followed by INR, severe TR status, cumulative in-hospital length-of-stay, cumulative number of hospital admissions, creatinine, urate, ALP, QTc and QT intervals.

Five-fold cross validation experiments were conducted on the dataset using GBST, RSF, and multivariate Cox regression (**Supplementary Table 3**). GBST showed the best survival prediction performance over RSF and multivariate Cox model according to evaluation metrics of precision, recall, AUC, C-Index, and KS-Index. The advantage of the GBST model stems from its ability to sequentially add weak decision tree learning models to the ensemble model, which can correct the prediction errors of prior models to minimize the overall prediction error. The predicted out-of-bag (OOB) survivals and cumulative hazards with the introduced GBST model are shown in **Supplementary Figure 3**.

## Discussion

The main findings of this territory-wide study of patients undergoing TAVR are two-fold. Firstly, patient demographics, comorbidities, healthcare utilization statistics prior to the procedure, laboratory examinations and ECG measurements were significant predictors of mortality. Secondly, a nonparametric gradient boosting survival tree model outperformed random survival forest model and multivariate Cox regression model for all-cause mortality prediction.

Frailty has been shown to be a strong predictor of adverse outcomes in patients with heart failure, and in those undergoing cardiac interventional procedures ^14-20^. Specifically related to TAVR, previous studies have examined the value of determining frailty for risk stratification. For example, a study in 2018 showed that 11% patients of average age of 83 died 2 years after TAVR, and a geriatric assessment frailty score cut-off at ≥4 predicted 2-year mortality with a specificity of 80% ^21^. Another study showed that 242 out of 544 TAVR patients were frail 1 year after the procedure based on frailty definition ^22^.

However, in clinical situations, it may be impractical to fully assess frailty status of patients and surrogates that can accurately model or reflect frailty would save time for patient assessment. Therefore, clinician-researchers have designed the electronic frailty index based on the concept that frailty is caused by the accumulation of health deficits ^23^. Segal *et al*. designed the electronic frailty index by selecting candidate variables based on their potential correlations with frailty state rather than mortality directly ^24^. An electronic frailty index has been used as an efficient variable to predict mortality in TAVR ^25^. In our territory-wide cohort, we developed an electronic frailty index based on predictors that were identified by Cox regression analysis and showed that this can predict all-cause mortality in patients undergoing TAVR.

Survival analysis has been widely used in clinical and epidemiological studies based on electronic health records (EHRs) that provide rich and diverse information for modelling and prediction. Survival analysis models in the literature may be parametric or semiparametric, including survival tree analysis in the context of conditional inference trees ^26^, survival forest analysis considering inverse probability of censoring weighting to compensate censoring ^27^, random survival forest model with log-rank test ^12^, censoring unbiased regression trees and forests with censoring unbiased loss functions ^28^, ensemble tree method for right-censored survival data ^29^. Traditional Cox proportional hazard models are used to identify the linear combinations. Tree structure-based survival analysis models have been widely applied in medical studies such as mortality prediction in systolic heart failure ^30^. In our study, we demonstrated that a nonparametric gradient boosting survival tree model significantly improved mortality prediction in patients undergoing TAVR.

## Conclusions

An electronic frailty index incorporating multi-domain data can efficiently predict all-cause mortality in patients undergoing TAVR. A machine learning-drive survival model significantly improves the risk prediction performance of the frailty models.

## Data Availability

The dataset has already been deposited in a repository and is accessible via: https://zenodo.org/record/4387098

## Acknowledgements

None.

## Funding

None.

## Data availability

The data underlying this article will be shared on reasonable request to the corresponding author.

## Conflicts of Interest

All authors declare no conflict of interest.

## Notes

### Competing Interest Statement

The authors have declared no competing interest.

### Author Declarations

This study was approved by The Joint Chinese University of Hong Kong - New Territories East Cluster Clinical Research Ethics Committee.

